# Hypertension Among Adults With HIV Initiating Antiretroviral Therapy in Freetown, Sierra Leone: A Cross-sectional Study

**DOI:** 10.1101/2023.06.17.23291458

**Authors:** George A. Yendewa, Darlinda F. Jiba, Daniel Sesay, Enanga Sonia Namanga, Sahr A. Yendewa, Umu Barrie, Gibrilla F. Deen, Foday Sahr, Robert A. Salata, James B.W. Russel, Sulaiman Lakoh

## Abstract

**Background:** Hypertension is a major contributor to cardiovascular morbidity and mortality in people with HIV (PWH). However, screening and identification among PWH initiating antiretroviral therapy (ART) in sub-Saharan Africa is understudied.We aimed to assess the prevalence of hypertension and its associated factors among newly diagnosed PWH in Freetown, Sierra Leone.

**Methods:** We used a cross-sectional study design to assess the baseline characteristics of newly diagnosed ART-naïve PWH at Connaught Hospital in Freetown from March 2021 to March 2022. We categorized patients as having normal blood pressure (<120/80mmHg), pre-hypertension (systolic 120-139 mmHg or diastolic 80-89 mmHg), and hypertension (systolic ≥140 mmHg or diastolic ≥90 mmHg). We used logistic regression models to identify factors independently associated with hypertension.

**Results:** 918 PWH were studied (55.0% female, median age 33 years). The prevalence of hypertension was 20.0% and 29.5% were pre-hypertensive. In univariate analysis, hypertension prevalence increased with age, body mass index (BMI), smoking, alcohol consumption, and being Christian (all p < 0.05). In multivariate logistic regression analysis, ages 40-49 years (aOR 2.15, 95% CI 1.30-3.57), 50-59 years (aOR 2.30, 95% CI 1.26-4.18), ≥ 60 years (aOR 3.08, 95% CI 1.28-7.41), BMI ≥ 30 kg/m^2^ (aOR 2.34, 95% CI 1.11-4.93), identifying as a Christian (aOR 1.45, 95% CI 1.01-2.11), and smoking (aOR 1.67, 95% CI 1.04-2.69) were significantly associated with hypertension.

**Conclusions:** We observed a significant burden of hypertension among ART-naïve PWH in Sierra Leone, emphasizing the importance of early identification and management to help reduce cardiovascular morbidity and mortality.

## BACKGROUND

Hypertension poses a significant public health problem, affecting an estimated 1.3 billion adults aged 30-79 years globally in 2021 [1]. The prevalence of hypertension is highest in the World Health Organization (WHO) African region, where approximately 27% of adults are hypertensive [1]. Despite the high global burden of the disease, less than half (42%) of individuals with hypertension are diagnosed and only 20% of those diagnosed are adequately treated, indicating a critical gap in awareness, detection and management [1]. Moreover, in 2021, hypertension remained the leading modifiable risk factor for premature cardiovascular disease (CVD)-related deaths, contributing to 11.3 million deaths globally and accounting for 2,770 per 100,000 disability-adjusted life years [2].

Recent evidence suggests an increasing prevalence of hypertension among people living with HIV (PWH) in comparison to the general population. This could be attributed to various factors, including HIV-induced chronic inflammation and immune dysfunction [3], toxicities associated with antiretroviral therapy (ART) [4], and rising rates of traditional cardiovascular risk factors such as aging, obesity, and lifestyle behaviors [6]. According to a 2017 systematic review and meta-analysis by Xu *et al* [6], about 35% of adult PWH on antiretroviral therapy (ART) and 13% of ART-naïve PWH globally had hypertension, in contrast to an estimated 30% among HIV-uninfected individuals. Additionally, several observational and modeling studies have forecasted an increase in hypertension rates in the coming decades [7, 8]. The projected rise in the burden of hypertension and other CVDs in low-income countries is driven by factors such as physical inactivity, dietary changes, work-related stress, and other unhealthy lifestyles associated with rapid urbanization, thus necessitating a focus on prevention, early detection, and management strategies [1, 9].

Sierra Leone, like many other countries in the WHO African region, is grappling with an increasing burden of HIV and CVDs risk factors [10-14]. While the national prevalence of HIV is estimated at 1.7% [10, 11], multiple studies have reported a high prevalence of hypertension in the general population. A recent population survey conducted by Russell *et al* [12] characterizing CVD risk factors in an urban setting in Sierra Leone reported a hypertension prevalence of 35.4% among adults. These findings are in concurrence with another study reporting a hypertension prevalence of 37.2% among healthcare workers [15], as well as another study among rural residents in Sierra Leone which reported a hypertension prevalence of 49.6% [13]. However, little attention has been given to the co-occurrence of hypertension among PWH in Sierra Leone. Recently, we reported a hypertension prevalence of 13% in a small cohort of ART-naïve patients in Sierra Leone (n=275), however, we did not access the correlates of hypertension [16].

The objective of the present study was to assess the prevalence of hypertension and its associated factors among ART-naïve PWH in Freetown, Sierra Leone. We specifically focused on individuals who were about to initiate ART as they represent a unique group at the early stage of HIV treatment and are likely to benefit from early interventions to prevent or manage hypertension.

## METHODS

### Study setting, design and population

We conducted a cross-sectional study of PWH enrolled in the Sierra Leone HIV Cohort Study at the HIV Clinic at Connaught Hospital in Freetown, Sierra Leone. The HIV Clinic at Connaught Hospital is the largest HIV treatment center in Sierra Leone, with over 4000 PWH in active clinical follow up. The Sierra Leone HIV Cohort Study was a prospective cohort study from 2020-2022 which consecutively enrolled individuals newly diagnosed with HIV, with the primary objective of evaluating their immunologic and virologic outcomes of newly diagnosed PWH who were initiated on ART in Sierra Leone. In the present cross-sectional study, we analyzed baseline sociodemographic and clinical data of adults aged ≥ 18 years enrolled from March 2021 to March 2022.

### Procedures and definitions

We collected baseline sociodemographic and clinical data of participants upon enrollment in the Sierra Leone HIV Cohort Study. Data points collected included age, sex, weight, height, marital status, residence (urban vs rural), highest level of education attained (none, primary, secondary or tertiary), religion (Christian vs Muslim), socioeconomic status (classified as low, middle or high), history of smoking, alcohol use, diabetic status, and blood pressure. For HIV-specific factors, we collected WHO staging at HIV diagnosis, as CD4 count and HIV viral load were not routine done. We stored the data in a password-protected spreadsheet accessible only to research team members. We removed all patient identifiers including name and personal identification numbers to ensure confidentiality.

Blood pressure was assessed according to the Eighth Joint National Committee (JNC8) guidelines [17] as the average of two readings or being on antihypertensive and classified as follows: (1) normal: systolic blood pressure (SBP) < 120 mmHg and diastolic blood pressure (DBP) < 80 mmHg; (2) pre-hypertension: SBP 120-139 mmHg or DBP 80-89 mmHg; and (3) hypertension: SBP ≥ 140 mmHg or DBP ≥ 90 mmHg. To ensure accurate BP measurements, participants were instructed to adopt a seated position, resting for a minimum of 5 minutes, with their left arm placed on a table and their legs uncrossed. To optimize accuracy, cuff size was selected based on mid-upper arm circumference.

Body mass index (BMI) was classified according to the United States Centers for Disease Control and Prevention criteria [18] as follows (1) underweight: < 18.5 kg/m^2^, (2) normal: 18.5-24.9 kg/m^2^, (3) overweight: 25-29.9 kg/m^2^, and obese: ≥ 30 kg/m^2^ for obesity. Diabetes was defined in accordance with the American Diabetic Association guidelines [19] as fasting glucose ≥ 126 mg/dL, 2-hour glucose ≥ 200 mg/dL, documented history of diabetes, or receiving antidiabetic medication. Smoking was regarded as any amount of tobacco smoking within the last 24 hours. Alcohol use was regarded as consuming > 20 g or 2 drinks of an alcoholic beverage daily. For the purpose of this study, we categorized socioeconomic status based on our knowledge of local economic conditions as low (earning < 120 USD/month), middle (earning 120-500 USD/month) and high (earning > 500 USD/month).

### Statistical analysis

We used the software SPSS Version 29.0 (Armonk, NY; IBM Corp) to perform the statistical analyses. Categorical variables were reported as frequencies and percentages and associations were assessed using Pearson’s chi-square test. Continuous variables were reported as medians and interquartile ranges (IQR) and associations assessed using the non-parametric independent samples Mann–Whitney U-test. Logistic regression models were used to identify associations between hypertension and covariates (age, sex, BMI, smoking, alcohol use, diabetes, immune status as defined by WHO stages) and potential confounders (relationship status, educational attainment, religion, socioeconomic status, and residence). Variables which attained a p-value of < 0.20 in the univariate analysis were included in the multivariable regression model. Adjusted odds ratios (aOR) and their corresponding 95% confidence intervals (CI) were calculated to estimate the strength of associations after controlling for potential confounders. In all calculations, statistical significance was set at p < 0.05.

### Ethical Approval

Ethical approval was obtained from the Sierra Leone Ethics and Scientific Review Committee (approved 9 March 2021). All study procedures were conducted in accordance with the ethical standards outlined in the Declaration of Helsinki. Informed consent was obtained from all study participants prior to their inclusion in the study.

## RESULTS

### Descriptive statistics of study population

Table 1 displays the characteristics of the study population. A total of 918 ART-naïve PWH were studied, of whom 505 (55.0%) were female. The distribution of participants across WHO stages at diagnosis was as follows: stage 1 (27.8%), stage 2 (25.7%), stage 3 (21.1%), and stage 4 (25.4%).

**Table 1.**
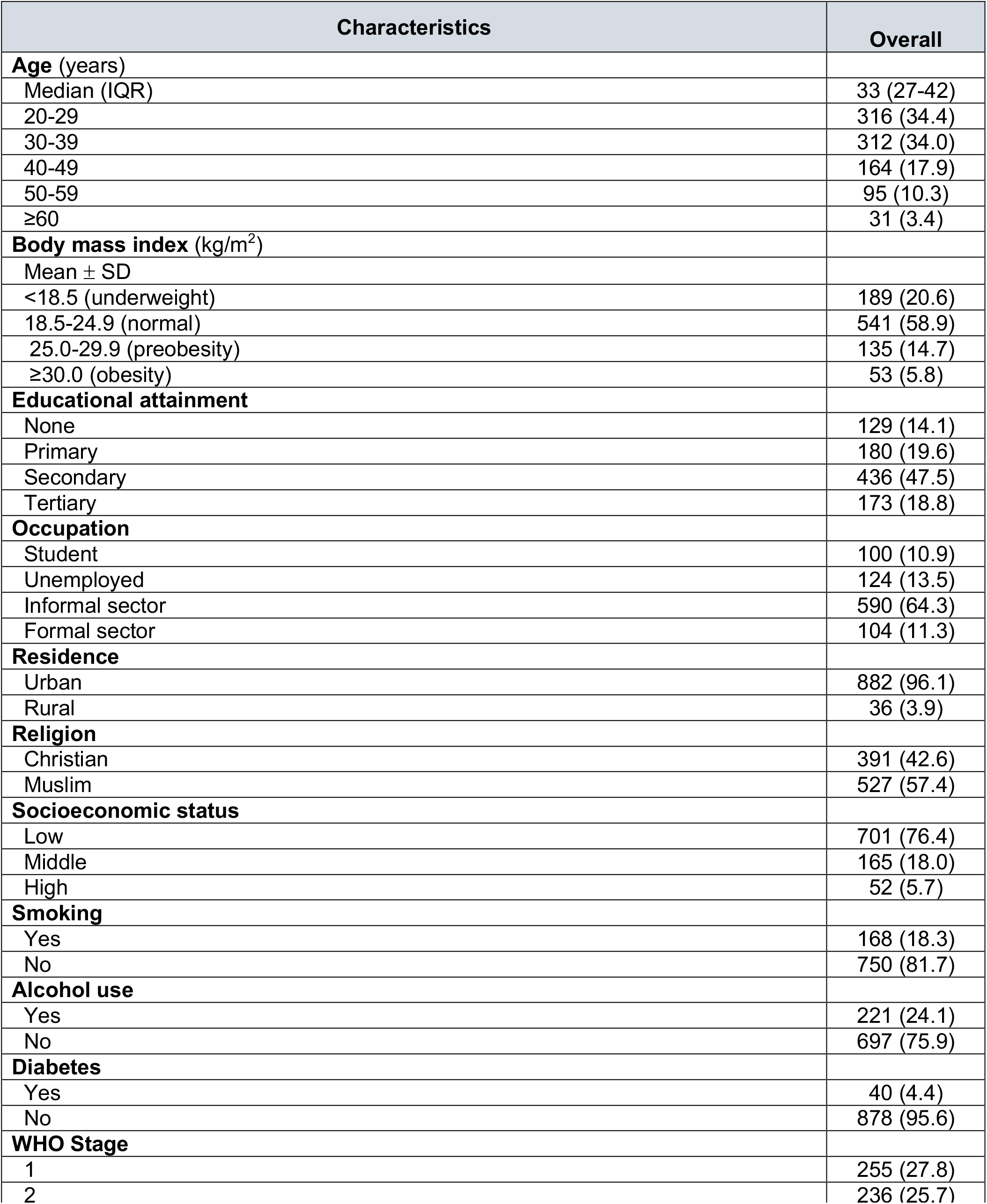
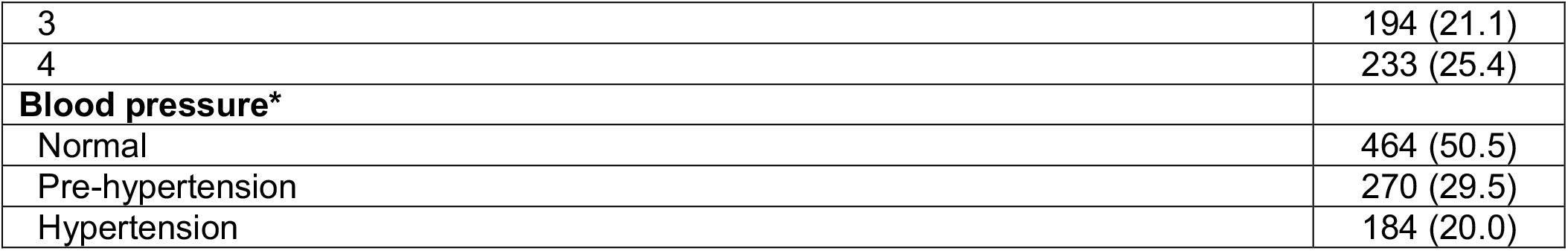
Characteristics of study participants stratified by hypertension status (N=918)

| Characteristics | Overall |
| --- | --- |
| **Age (years)** |  |
| Median (IQR) | 33 (27-42) |
| 20-29 | 316 (34.4) |
| 30-39 | 312 (34.0) |
| 40-49 | 164 (17.9) |
| 50-59 | 95 (10.3) |
| ≥60 | 31 (3.4) |
| **Body mass index (kg/m<sup>2</sup>)** |  |
| Mean ± SD |  |
| <18.5 (underweight) | 189 (20.6) |
| 18.5-24.9 (normal) | 541 (58.9) |
| 25.0-29.9 (preobesity) | 135 (14.7) |
| ≥30.0 (obesity) | 53 (5.8) |
| **Educational attainment** |  |
| None | 129 (14.1) |
| Primary | 180 (19.6) |
| Secondary | 436 (47.5) |
| Tertiary | 173 (18.8) |
| **Occupation** |  |
| Student | 100 (10.9) |
| Unemployed | 124 (13.5) |
| Informal sector | 590 (64.3) |
| Formal sector | 104 (11.3) |
| **Residence** |  |
| Urban | 882 (96.1) |
| Rural | 36 (3.9) |
| **Religion** |  |
| Christian | 391 (42.6) |
| Muslim | 527 (57.4) |
| **Socioeconomic status** |  |
| Low | 701 (76.4) |
| Middle | 165 (18.0) |
| High | 52 (5.7) |
| **Smoking** |  |
| Yes | 168 (18.3) |
| No | 750 (81.7) |
| **Alcohol use** |  |
| Yes | 221 (24.1) |
| No | 697 (75.9) |
| **Diabetes** |  |
| Yes | 40 (4.4) |
| No | 878 (95.6) |
| **WHO Stage** |  |
| 1 | 255 (27.8) |
| 2 | 236 (25.7) |
| 3 | 194 (21.1) |
| 4 | 233 (25.4) |
| **Blood pressure\*** |  |
| Normal | 464 (50.5) |
| Pre-hypertension | 270 (29.5) |
| Hypertension | 184 (20.0) |

The median age was 33 years (IQR 27-42). The median BMI was 22 kg/m^2^ (IQR 19-24), with 188 (20.5%) classified as overweight or obese. In terms of educational attainment, the majority had primary education or higher (818, 85.9%). Most participants were employed in the informal sector (64.3%), while 11.3% were engaged in the formal sector. The majority resided in urban areas (882, 96.1%). Regarding religious affiliation, 527 (57.4%) identified as Muslim, while 391 (42.6%) identified as Christian. In terms of lifestyle factors, 168 (18.3%) participants reported smoking, while 221 (24.1%) acknowledged regular alcohol consumption. The prevalence of diabetes among the participants was 4.4%.

### Prevalence of hypertension

Overall, 184 participants had hypertension, yielding a prevalence of 20.0%. Furthermore, 270 participants (29.4%) were classified as having pre-hypertension, while about half (464, 50.5%) had normal blood pressure (Table 1).

Table 2 displays the distribution of characteristics among PWH with and without hypertension. We observed no significant differences in hypertension prevalence between males and females (p=0.840), urban vs rural residence (p=0.108), diabetic status (p=0.692), or WHO Stage at diagnosis (p=0.086). However, the prevalence of hypertension varied with increasing age (p<0.001) and increasing BMI (p=0.043). Among sociodemographic factors, having secondary education (42.4%, p=0.027), being of middle socioeconomic status (25.5%, p=0.002), and being Christian (52.7%, p=0.002) were associated with hypertension. Additionally, hypertension was significantly more common with smoking (26.6%, p=0.001) and alcohol use (33.7%; p<0.001).

**Table 2.** Comparison of PHW with and without hypertension

| Characteristics | Hypertension |  | p-Value |
| --- | --- | --- | --- |
|  | Yes | No |  |
| **Sex** |  |  |  |
| Male | 84 (45.7) | 329 (44.8) | 0.840 |
| Female | 100 (54.3) | 405 (55.2) |  |
| **Age (years)** |  |  |  |
| 20-29 | 42 (22.8) | 274 (37.3) | **<0.001** |
| 30-39 | 56 (30.4) | 256 (34.9) |  |
| 40-49 | 48 (26.1) | 116 (15.8) |  |
| 50-59 | 28 (15.2) | 67 (9.1) |  |
| ≥60 | 10 (5.4) | 21 (2.9) |  |
| **Body mass index (kg/m<sup>2</sup>)** |  |  |  |
| <18.5 (underweight) | 28 (15.2) | 161 (21.9) | **0.043** |
| 18.5-24.9 (normal) | 111 (60.3) | 430 (58.6) |  |
| 25.0-29.9 (preobesity) | 28 (15.2) | 107 (14.6) |  |
| ≥30.0 (obesity) | 17 (9.2) | 36 (4.9) |  |
| **Educational attainment** |  |  |  |
| None | 27 (14.7) | 102 (13.9) | **0.027** |
| Primary | 50 (27.2) | 130 (17.7) |  |
| Secondary | 78 (42.4) | 358 (48.8) |  |
| Tertiary | 29 (15.8) | 144 (19.6) |  |
| **Occupation** |  |  |  |
| Student | 10 (5.4) | 90 (12.3) | **0.034** |
| Unemployed | 23 (12.5) | 101 (13.8) |  |
| Informal sector | 132 (71.7) | 458 (62.4) |  |
| Formal sector | 19 (10.3) | 85 (11.6) |  |
| **Residence** |  |  |  |
| Urban | 173 (94.0) | 709 (96.6) | 0.108 |
| Rural | 11 (6.0) | 25 (3.4) |  |
| **Religion** |  |  |  |
| Christian | 97 (52.7) | 294 (40.1) | **0.002** |
| Muslim | 87 (47.3) | 440 (59.9) |  |
| **Socioeconomic status** |  |  |  |
| Low | 129 (70.1) | 572 (77.9) | **0.010** |
| Middle | 47 (25.5) | 118 (16.1) |  |
| High | 8 (4.3) | 44 (6.0) |  |
| **Smoking** |  |  |  |
| Yes | 49 (26.6) | 119 (16.2) | **0.001** |
| No | 135 (73.4) | 615 (83.8) |  |
| **Alcohol use** |  |  |  |
| Yes | 62 (33.7) | 159 (21.7) | **<0.001** |
| No | 122 (66.3) | 575 (78.3) |  |
| **Diabetes** |  |  |  |
| Yes | 9 (4.9) | 31 (4.2) | 0.692 |
| No | 175 (95.1) | 703 (95.8) |  |
| **WHO Stage** |  |  |  |
| 1 | 54 (29.3) | 201 (27.4) | 0.086 |
| 2 | 48 (26.1) | 188 (25.6) |  |
| 3 | 27 (14.7) | 167 (22.8) |  |
| 4 | 55 (29.9) | 178 (24.3) |  |

### Factors associated with hypertension

Table 3 shows the results of the univariate and multivariate regression model. When adjusting for confounders, we observed that increasing age showed a significant association with hypertension, with higher odds for age groups 40-49 years (AOR 2.15, 95% CI [1.30-3.57]; p=0.003), 50-59 years (aOR 2.30, 95% CI [1.26-4.18]; p=0.006), and ≥ 60 years (aOR 3.08, 95% CI [1.28-7.41]; p=0.012). Additionally, obesity (aOR 2.34, 95% CI [1.11-4.93]; p=0.025), identifying as Christian (aOR 1.45, 95% CI [1.01-2.11]; p=0.047), and smoking (aOR 1.67, 95% CI [1.04-2.69]; p=0.033) were significantly associated with hypertension.

**Table 3.** Univariate and multivariate analyses of factors associated with hypertension

| Characteristics | Univariate |  | Multivariate |  |
| --- | --- | --- | --- | --- |
|  | Crude OR (95% CI) | p-Value | Adjusted OR (95% CI) | p-Value |
| **Sex** |  |  |  |  |
| Male | 1.03 (0.75-1.43) | 0.840 |  |  |
| Female | Ref |  |  |  |
| **Age (years)** |  |  |  |  |
| 20-29 | Ref |  | Ref |  |
| 30-39 | 1.43 (0.92-2.20) | 0.109 | 1.27 (0.80-2.00) | 0.304 |
| 40-49 | 2.70 (1.69-4.31) | **<0.001** | 2.15 (1.30-3.57) | **0.003** |
| 50-59 | 2.72 (1.58-4.72) | **<0.001** | 2.30 (1.26-4.18) | **0.006** |
| ≥60 | 3.11 (1.37-7.04) | **0.007** | 3.08 (1.28-7.41) | **0.012** |
| **Body mass index (kg/m<sup>2</sup>)** |  |  |  |  |
| <18.5 (underweight) | Ref |  | Ref |  |
| 18.5-24.9 (normal) | 1.48 (0.94-2.33) | 0.087 | 1.39 (0.86-2.23) | 0.180 |
| 25.0-29.9 (preobesity) | 1.50 (0.84-2.68) | 0.166 | 1.37 (0.74-2.53) | 0.311 |
| ≥30.0 (obesity) | 2.72 (1.34-5.49) | **0.005** | 2.34 (1.11-4.93) | **0.025** |
| **Educational attainment** |  |  |  |  |
| None | Ref |  | Ref |  |
| Primary | 1.45 (0.85-2.48) | 0.171 | 1.10 (0.61-1.99) | 0.108 |
| Secondary | 0.82 (0.50-1.34) | 0.436 | 0.82 (0.48-1.39) | 0.540 |
| Tertiary | 0.76 (0.42-1.36) | 0.357 | 0.83 (0.41-1.66) | 0.286 |
| **Occupation** |  |  |  |  |
| Student | Ref |  | Ref |  |
| Unemployed | 2.05 (0.93-4.55) | 0.077 | 1.27 (0.52-3.12) | 0.599 |
| Informal sector | 2.59 (1.31-5.13) | **0.006** | 1.57 (0.74-3.33) | 0.244 |
| Formal sector | 2.00 (0.88-4.57) | 0.095 | 1.16 (0.48-2.85) | 0.738 |
| **Residence** |  |  |  |  |
| Urban | 0.55 (0.27-1.15) | 0.108 | 0.52 (0.24-1.14) | 0.100 |
| Rural | Ref |  | Ref |  |
| **Religion** |  |  |  |  |
| Christian | 1.67 (1.21-2.31) | **0.002** | 1.45 (1.01-2.11) | **0.047** |
| Muslim | Ref |  | Ref |  |
| **Socioeconomic status** |  |  |  |  |
| Low | 1.77 (1.20-2.60) | **0.004** | 1.52 (0.94-2.44) | 0.086 |
| Middle | 0.81 (0.37-1.75) | 0.587 | 0.71 (0.30-1.69) | 0.434 |
| High | Ref |  | Ref |  |
| **Smoking** |  |  |  |  |
| Yes | 1.88 (1.28-2.75) | **0.001** | 1.67 (1.04-2.69) | **0.033** |
| No | Ref |  | Ref |  |
| **Alcohol use** |  |  |  |  |
| Yes | 1.84 (1.29-2.62) | **<0.001** | 1.30 (0.82-2.04) | 0.262 |
| No | Ref |  | Ref |  |
| **Diabetes** |  |  |  |  |
| Yes | 1.17 (0.55-2.49) | 0.692 |  |  |
| No | Ref |  |  |  |
| **WHO Stage** |  |  |  |  |
| 1 | Ref |  | Ref |  |
| 2 | 0.95 (0.61-1.47) | 0.819 | 0.83 (0.52-1.31) | 0.420 |
| 3 | 0.60 (0.36-1.00) | **0.049** | 0.53 (0.31-0.92) | **0.023** |
| 4 | 1.15 (0.75-1.76) | 0.520 | 1.02 (0.57-1.67) | 0.936 |

## DISCUSSION

In this study, we observed that 20.0% of PWH who were initiating ART at Connaught Hospital in Freetown, Sierra Leone had hypertension. Additionally, 29.4% had pre-hypertension, indicating a concerning trend, considering that the median age of this cohort of patients was only 33 years. Our findings support current research from the WHO African region which has shown substantially high rates of hypertension among both ART-naïve and -exposed PWH [20-23]. The findings of this study contribute to the limited literature on hypertension among PWH in Sierra Leone specifically and may provide valuable insights for HIV care providers and policymakers in designing appropriate interventions to address the growing burden of hypertension and CVDs in this vulnerable population.

We identified sociodemographic and lifestyle-associated factors as the primary determinants of hypertension in our study. There was a strong association between hypertension and increasing age, with a 2-to 3-fold higher odds in prevalence observed in PWH aged 30 years or older. Additionally, about 1 in 5 (20.5%) were either overweight or obese, with obesity specifically conferring a 2.34-fold high odds of having hypertension. Furthermore, a similar proportion (18.8%) were smokers, which increased their odds of hypertension 1.67-fold. These findings are consistent with the shifting epidemiological trends in sub-Saharan Africa, where the increasing prevalence of classic CVD risk factors has significantly contributed to the rise in hypertension and other CVDs in this region [1, 24]. To improve cardiovascular health outcomes, HIV programs in SSA must integrate NCD services into routine HIV care, focusing on modifiable risk factors including promoting healthy behaviors such as regular physical activity, healthy diets, and smoking cessation [24, 25].

Other sociodemographic factors that are commonly associated with hypertension include alcohol use, educational status, occupation, and urban versus rural residence [26, 27]. Generally, higher education, better employment, and urban residence are indicators of higher socioeconomic status, which correlates with increased health literacy, better knowledge of disease processes, and better access to healthcare services [26, 27]. Urban dwelling may also be linked to unhealthy behaviors such as increased smoking, higher alcohol consumption, unhealthy dietary habits, sedentary lifestyles, and greater exposure to environmental pollutants, which can negatively impact cardiovascular health [27 28]. However, our study did not find these sociodemographic factors to be independently associated with hypertension. Several plausible reasons can explain this. For instance, the majority (85.9%) of participants had primary education or higher, creating a skewed distribution that favored educated individuals over those with lower education. Similarly, our cohort comprised only 4% of individuals residing in rural areas, limiting the ability to compare the prevalence of hypertension between urban and rural residents. This highlights the need for larger and more representative samples to comprehensively examine the relationship between sociodemographic factors and hypertension.

Interestingly, individuals identifying as Muslims exhibited a 1.45-fold lower odds of hypertension compared to Christians. Religious practices and beliefs, including fasting, prayer, meditation, and mindfulness, have been investigated for their impact on blood pressure and other health outcomes. A meta-analysis by Al-Jafar et al [29] involving 33 studies and 3213 Muslim participants revealed a decrease in systolic blood pressure by 3.19 mm Hg and diastolic blood pressure by 2.26 mm Hg after Ramadan fasting, a significant Islamic practice. These improvements were observed in both healthy individuals and those with hypertension or diabetes [29]. Nonetheless, it is essential to acknowledge that blood pressure is influenced by multiple factors such as genetics, lifestyle, diet, physical activity, and overall health, which can differ among individuals within and from different religious groups [29, 30]. Further research considering various sociodemographic, lifestyle, and health-related factors among individuals of diverse religious backgrounds is necessary to gain a comprehensive understanding of the relationship between religious affiliation and blood pressure.

The prevalence of diabetes in our study was 4.4%, in agreement with previous research on the HIV and general populations in Sierra Leone, which have reported prevalence rates ranging from 2.4% to 8.9% [12, 13, 31, 32]. However, it is important to acknowledge that diabetes is often underdiagnosed, especially among people living with HIV [33]. Diabetes, along with hypertension are major components of the metabolic syndrome, which is a significant CVD risk factor [2, 34]. The rising incidence of diabetes, hypertension, and the metabolic syndrome presents substantial challenges in low-income settings, especially in sub-Saharan Africa [35]. This problem is likely to be exacerbated even further by the introduction of dolutegravir as first-line ART, which has been associated with weight gain, insulin resistance, and metabolic complications [4, 36]. Therefore, the integration of diabetes screening and comprehensive care into HIV programs is of utmost importance to address the mounting burden of diabetes and its implications for PWH in the era of dolutegravir and other newer-generation integrase strand transfer inhibitors.

We acknowledge the limitations of our study. Firstly, the cross-sectional design limits the ability to establish causal relationships between the identified factors and hypertension; longitudinal studies would provide a better understanding of the associations and potential mechanisms underlying these relationships. Secondly, this was a single-center study from an urban setting, which limits its generalizability. Thirdly, we were unable to investigate the impact of other cardiovascular correlates such as immune status as determined by CD4 count, HIV viremia, level of physical activity, plasma lipid profile, and dietary habits such as salt intake, and family history, all of which are known determinants of hypertension risk. Additionally, the reliance on self-reported lifestyle factors may introduce recall bias and misclassification. Nevertheless, our study highlights the increasing burden of hypertension among PWH in Africa and underscores the importance of early screening to facilitate timely interventions to reduce future cardiovascular complications.

## CONCLUSION

In summary, our study found a prevalence of 20% of hypertension and a pre-hypertension prevalence of 29.4% in a large cohort of newly diagnosed PWH prior to initiating ART in Freetown, Sierra Leone. Older age, obesity, smoking, and identifying as a Christian were independent predictors of hypertension. These findings underscore the need for comprehensive cardiovascular risk assessment and tailored interventions targeting modifiable risk factors at ART initiation to effectively prevent and manage hypertension among PWH in Sierra Leone and other low-income settings in SSA.

## Data Availability

All data produced in the present work are contained in the manuscript

## AUTHOR CONTRIBUTIONS

GAY, RAS, JBWR and SL conceptualized the study. DFJ, DS, ESN, SAY and UB collected the data. GAY conducted the statistical analysis. GAY wrote the initial manuscript draft. DFJ, DS, ESN, SAY, UB, GFD, and FS participated in critically reviewing the manuscript. All authors contributed intellectual content and approved he final version. GAY is acting as the guarantor of this manuscript.

## DATA AVAILABILITY STATEMENT

The data presented in this study are contained in the manuscript.

## REFERENCES

1. World Health Organization 2023. Hypertension. Available at: https://www.who.int/news-room/fact-sheets/detail/hypertension. xAccessed on June 15, 2023.

2. Razo C, Welgan CA, Johnson CO, McLaughlin SA, Iannucci V, Rodgers A, Wang N, LeGrand KE, Sorensen RJD, He J, Zheng P, Aravkin AY, Hay SI, Murray CJL, Roth GA. Effects of elevated systolic blood pressure on ischemic heart disease: a Burden of Proof study. Nat Med. 2022;28(10):2056–2065.

3. Currier JS, Lundgren JD, Carr A, Klein D, Sabin CA, Sax PE, Schouten JT, Smieja M; Working Group 2. Epidemiological evidence for cardiovascular disease in HIV-infected patients and relationship to highly active antiretroviral therapy. Circulation. 2008;118(2):e29–35.

4. Sax PE, Erlandson KM, Lake JE, Mccomsey GA, Orkin C, Esser S, Brown TT, Rockstroh JK, Wei X, Carter CC, Zhong L, Brainard DM, Melbourne K, Das M, Stellbrink HJ, Post FA, Waters L, Koethe JR. Weight Gain Following Initiation of Antiretroviral Therapy: Risk Factors in Randomized Comparative Clinical Trials. Clin Infect Dis. 2020 Sep 12;71(6):1379–1389.

5. So-Armah K, Benjamin LA, Bloomfield GS, Feinstein MJ, Hsue P, Njuguna B, Freiberg MS. HIV and cardiovascular disease. Lancet HIV. 2020;7(4):e279–e293.

6. Xu Y, Chen X, Wang K. Global prevalence of hypertension among people living with HIV: a systematic review and meta-analysis. J Am Soc Hypertens. 2017;11(8):530–540.

7. Bosu WK, Reilly ST, Aheto JMK, Zucchelli E. Hypertension in older adults in Africa: A systematic review and meta-analysis. PLoS One. 2019;14(4):e0214934.

8. Smit M, Brinkman K, Geerlings S, Smit C, Thyagarajan K, Sighem Av, de Wolf F, Hallett TB; ATHENA observational cohort. Future challenges for clinical care of an ageing population infected with HIV: a modelling study. Lancet Infect Dis. 2015;15(7):810–8.

9. UNAIDS 2021. Responding to the Challenge of Non-communicable Diseases. Available at: https://www.unaids.org/sites/default/files/media_asset/responding-to-the-challenge-of-non-communicable-diseases_en.pdf. xAccessed on June 15, 2023.

10. UNAIDS 2020. Country progress report-Sierra Leone. Available at: https://www.unaids.org/sites/default/files/country/documents/SLE_2020_countryreport.pdf Accessed on June 15, 2023.

11. Yendewa GA, Poveda E, Yendewa SA, Sahr F, Quiñones-Mateu ME, Salata RA. HIV/AIDS in Sierra Leone: Characterizing the Hidden Epidemic. AIDS Rev. 2018;20(2):104–113.

12. Russell JBW, Koroma TR, Sesay S, Samura SK, Lakoh S, Bockarie A, Abir OT, Kanu JS, Coker J, Jalloh A, Conteh V, Conteh S, Smith M, Mahdi OZ, Lisk DR. Burden of cardiometabolic risk factors and preclinical target organ damage among adults in Freetown, Sierra Leone: a community-based health-screening survey. BMJ Open. 2023;13(5):e067643.

13. Odland ML, Bockarie T, Wurie H, Ansumana R, Lamin J, Nugent R, Bakolis I, Witham M, Davies J. Prevalence and access to care for cardiovascular risk factors in older people in Sierra Leone: a cross-sectional survey. BMJ Open. 2020;10(9):e038520.

14. Bockarie T, Odland ML, Wurie H, Ansumana R, Lamin J, Witham M, Oyebode O, Davies J. Prevalence and socio-demographic associations of diet and physical activity risk-factors for cardiovascular disease in Bo, Sierra Leone. BMC Public Health. 2021;21(1):1530.

15. Russell JB, Rahman-Sesay J, Conteh V, Conteh S, Jalloh AP, Ibrahim-Sayo E, Lisk DR. Prevalence, Awareness and Risk Factors of Hypertension among Health Workers at the Connaught Teaching Hospital, Sierra Leone. West Afr J Med. 2020;37(5):450–459.

16. Yendewa GA, Lakoh S, Jiba DF, Yendewa SA, Barrie U, Deen GF, Samai M, Jacobson JM, Sahr F, Salata RA. Hepatitis B Virus and Tuberculosis Are Associated with Increased Noncommunicable Disease Risk among Treatment-Naïve People with HIV: Opportunities for Prevention, Early Detection and Management of Comorbidities in Sierra Leone. J Clin Med. 2022;11(12):3466.

17. James PA, Oparil S, Carter BL, Cushman WC, Dennison-Himmelfarb C, Handler J, Lackland DT, LeFevre ML, MacKenzie TD, Ogedegbe O, Smith SC Jr, Svetkey LP, Taler SJ, Townsend RR, Wright JT Jr, Narva AS, Ortiz E. 2014 evidence-based guideline for the management of high blood pressure in adults: report from the panel members appointed to the Eighth Joint National Committee (JNC 8). JAMA. 2014;311(5):507–20.

18. Centers for Disease Control and Prevention. About Adult BMI-Healthy Weight, Nutrition, and Physical Activity. Available at: https://www.cdc.gov/healthyweight/assessing/bmi/adult_bmi/index.html Accessed on June 15, 2023.

19. American Diabetes Association. Diagnosis and classification of diabetes mellitus. Diabetes Care. 2014; 37 Suppl 1:S81–S90.

20. Migisha R, Ario AR, Kadobera D, Bulage L, Katana E, Ndyabakira A, Elyanu P, Kalamya JN, Harris JR. High blood pressure and associated factors among HIV-infected young persons aged 13 to 25 years at selected health facilities in Rwenzori region, western Uganda, September-October 2021. Clin Hypertens. 2023;29(1):6.

21. Amutuhaire W, Mulindwa F, Castelnuovo B, Brusselaers N, Schwarz JM, Edrisa M, Dujanga S, Salata RA, Yendewa GA. Prevalence of Cardiometabolic Disease Risk Factors in People with HIV Initiating Antiretroviral Therapy at a High-Volume HIV Clinic in Kampala, Uganda. Open Forum Infect Dis. 2023;10(5):ofad241.

22. Isaac Derick K, Khan Z. Prevalence, Awareness, Treatment, Control of Hypertension, and Availability of Hypertension Services for Patients Living With Human Immunodeficiency Virus (HIV) in Sub-Saharan Africa (SSA): A Systematic Review and Meta-analysis. Cureus. 2023;15(4):e37422.

23. Bigna JJ, Ndoadoumgue AL, Nansseu JR, Tochie JN, Nyaga UF, Nkeck JR, Foka AJ, Kaze AD, Noubiap JJ. Global burden of hypertension among people living with HIV in the era of increased life expectancy: a systematic review and meta-analysis. J Hypertens. 2020;38(9):1659–1668.

24. Minja NW, Nakagaayi D, Aliku T, Zhang W, Ssinabulya I, Nabaale J, Amutuhaire W, de Loizaga SR, Ndagire E, Rwebembera J, Okello E, Kayima J. Cardiovascular diseases in Africa in the twenty-first century: Gaps and priorities going forward. Front Cardiovasc Med. 2022;9:1008335.

25. Amegah AK. Tackling the Growing Burden of Cardiovascular Diseases in Sub-Saharan Africa. Circulation. 2018 Nov 27;138(22):2449–2451.

26. van Oort S, Beulens JWJ, van Ballegooijen AJ, Grobbee DE, Larsson SC. Association of Cardiovascular Risk Factors and Lifestyle Behaviors With Hypertension: A Mendelian Randomization Study. Hypertension. 2020;76(6):1971–1979.

27. Sani RN, Connelly PJ, Toft M, Rowa-Dewar N, Delles C, Gasevic D, Karaye KM. Rural-urban difference in the prevalence of hypertension in West Africa: a systematic review and meta-analysis. J Hum Hypertens. 2022;10.1038/s41371-022-00688-8.

28. Wang J, Sun W, Wells GA, Li Z, Li T, Wu J, Zhang Y, Liu Y, Li L, Yu Y, Liu Y, Qi C, Lu Y, Liu N, Yan Y, Liu L, Hui G, Liu B. Differences in prevalence of hypertension and associated risk factors in urban and rural residents of the northeastern region of the People’s Republic of China: A cross-sectional study. PLoS One. 2018;13(4):e0195340.

29. Al-Jafar R, Zografou Themeli M, Zaman S, Akbar S, Lhoste V, Khamliche A, Elliott P, Tsilidis KK, Dehghan A. Effect of Religious Fasting in Ramadan on Blood Pressure: Results From LORANS (London Ramadan Study) and a Meta-Analysis. J Am Heart Assoc. 2021;10(20):e021560.

30. Kawachi I. Invited Commentary: Religion as a Social Determinant of Health. Am J Epidemiol. 2020;189(12):1461–1463.

31. Ceesay MM, Morgan MW, Kamanda MO, Willoughby VR, Lisk DR. Prevalence of diabetes in rural and urban populations in southern Sierra Leone: a preliminary survey. Trop Med Int Health. 1997;2(3):272–277.

32. Sundufu AJ, Bockarie CN, Jacobsen KH. The prevalence of type 2 diabetes in urban Bo, Sierra Leone, and in the 16 countries of the West Africa region. Diabetes Metab Res Rev. 2017;33(7):10.1002/dmrr.2904.

33. Kousignian I, Sautereau A, Vigouroux C, Cros A, Kretz S, Viard JP, Slama L. Diagnosis, risk factors and management of diabetes mellitus in HIV-infected persons in France: A real-life setting study. PLoS One. 2021;16(5):e0250676.

34. Alberti KG, Eckel RH, Grundy SM, Zimmet PZ, Cleeman JI, Donato KA, Fruchart JC, James WP, Loria CM, Smith SC Jr; International Diabetes Federation Task Force on Epidemiology and Prevention; Hational Heart, Lung, and Blood Institute; American Heart Association; World Heart Federation; International Atherosclerosis Society; International Association for the Study of Obesity. Harmonizing the metabolic syndrome: a joint interim statement of the International Diabetes Federation Task Force on Epidemiology and Prevention; National Heart, Lung, and Blood Institute; American Heart Association; World Heart Federation; International Atherosclerosis Society; and International Association for the Study of Obesity. Circulation. 2009;120(16):1640–1645.

35. Jaspers Faijer-Westerink H, Kengne AP, Meeks KAC, Agyemang C. Prevalence of metabolic syndrome in sub-Saharan Africa: A systematic review and meta-analysis. Nutr Metab Cardiovasc Dis. 2020;30(4):547–565.

36. Eckard AR, McComsey GA. Weight gain and integrase inhibitors. Curr Opin Infect Dis. 2020;33(1):10–19.

